# Analytical Validation of Minimally Invasive Capillary Blood Microsampling using Tasso+ for Multiplexed Neurological Biomarkers

**DOI:** 10.64898/2026.05.15.26353201

**Authors:** Owen Swann, Sophie Hicks, Caleigh Lynch, Amie Wallman-Jones, Maryam Shoai, Rachel Mulvaney, Bárbara Fernandes Gomes, Eleftheria Kodosaki, Margherita Tecilla, Mazdak Ghajari, Ben Jones, Simon Kemp, TBI-REPORTER Biomarker Group, Richard Sylvester, Matt Cross, Keith Stokes, Mathew G Wilson, David K Menon, Amanda Heslegrave, Henrik Zetterberg, David J Sharp, Thomas D Parker

**Author notes:** **Corresponding author:** Owen Swann 3rd floor, Queen Square House, Institute of Neurology, Queen Square London WC1N 3BG. TBI-Reporter, https://tbi-reporter.uk/.

## Abstract

Blood-based biomarkers are increasingly used to investigate brain health, but collecting venous blood is difficult in remote and field settings. Capillary microsampling offers a practical alternative, although the ability to delay processing and its agreement with gold-standard venous blood require validation. We evaluated Tasso+, a minimally invasive upper-arm capillary blood collection system, for measuring neurological and host-response biomarkers in plasma and serum during an exercise-based protocol. Sampling occurred before, immediately after, and approximately 24-to-36 hours after exercise; Tasso+ samples were processed with or without a 72-hour room-temperature delay. Tasso+ samples were compared with matched venous blood, and Capitainer SEP10 dried plasma spots were also evaluated, using Quanterix Simoa 4plexD+ and Alamar Biosciences NULISAseq CNS panel. Tasso+ enabled reliable measurement of several key biomarkers, even after delayed processing. These findings support capillary microsampling for neurological biomarker studies where venepuncture is challenging, including field-based research and participant-led remote sampling.

## Introduction

Blood-based biomarkers are increasingly central to studying brain health across neurological and neurodegenerative conditions. They offer insight into pathological processes previously assessed via cerebrospinal fluid or neuroimaging^1,2^. Venous blood collection is the gold standard yet often impractical in non-clinical settings requiring rapid or repeated sampling^2^.

Recent work has explored remote finger-prick blood collection, with encouraging stability of neurofilament light chain (NfL) and glial fibrillary acidic protein (GFAP) after a 72 hour (h) processing delay^3^. However, this form of capillary sampling often requires manipulation (“milking”) to maintain blood flow^4^, which increases haemolysis^5^ and may bias ultrasensitive immunoassays^6^. Alternative capillary microsampling devices, including the Tasso+ device have emerged. Tasso+ is an upper-arm capillary blood collection device that adheres to the skin and uses a small lancet and gentle vacuum to collect blood into a detachable tube, enabling minimally invasive collection without conventional venepuncture. Tasso+ samples show low levels of haemolysis^5^, are compatible with highly multiplexed immunoassays^7^ and enable participant-led remote collection^8^ supporting scalable, repeat sampling in non-clinical settings. Understanding of biomarker behaviour following Tasso+ sampling is essential for its implementation.

Here, we evaluated the pre-analytical performance of Tasso+ and Capitainer SEP10 as complementary capillary microsampling approaches. Tasso+ collects liquid whole blood for subsequent plasma and serum processing, whereas Capitainer SEP10 separates plasma from whole blood on-card and stores dried plasma on collection discs intended for prolonged room-temperature stability. Tasso+ plasma and serum samples were processed without a delay (0 h) or with a delay (72 h), selected to reflect a realistic delay in sample processing following remote postal collection. Healthy adults were sampled at pre-exercise (baseline), immediately post-exercise and at a 24-to-36 h follow-up post-exercise, allowing assessment of exercise-associated biomarker changes and recovery dynamics; this is important in research contexts such as sport-related head impact studies, as exercise has been shown to influence some biomarkers relevant to neurological disease independently of brain injury ^9^.

Neurological blood-based biomarkers were assessed using Alamar Bioscience’s nucleic acid-linked immuno-sandwich assay (NULISAseq) central nervous system (CNS) disease panel 120 and the Quanterix single molecule array (SIMOA) Neurology 4-Plex D Plus assays. These assays are being widely used in neurological studies including neurodegeneration and traumatic brain injury (TBI). Primary analyses focused on agreement between Tasso+ and venous sampling for SIMOA-measured GFAP and NfL when processed without a delay (0 h) and with a delay (72 h). Secondary analyses extended these comparisons to the NULISAseq panel assessing 120 CNS relevant proteins. Tasso+ collection was also compared to Capitainer SEP10 dried samples collection. Exercise-related changes were evaluated as exploratory outcomes.

## Methods

### Study design and participants

Samples were collected at pre-exercise (baseline), immediately post-exercise and at a 24-to-36 h follow-up, yielding 168 samples per assay from 8 healthy adult participants. Participants provided written informed consent, completed the Physical Activity Readiness Questionnaire (PAR-Q), and underwent a resting 12-lead electrocardiogram to confirm eligibility for high-intensity exercise. Samples included 24 venous plasma, 24 venous serum, 24 Tasso+ plasma samples processed without a delay (0 h), 24 Tasso+ plasma samples processed with a delay (72 h), 24 Tasso+ serum samples processed without a delay (0 h), 24 Tasso+ serum samples processed with a delay (72 h) and 24 Capitainer SEP10 card samples. The study design, sample collection workflow, processing conditions and downstream analytical platforms are summarised in Figure 1.

**Figure 1.**
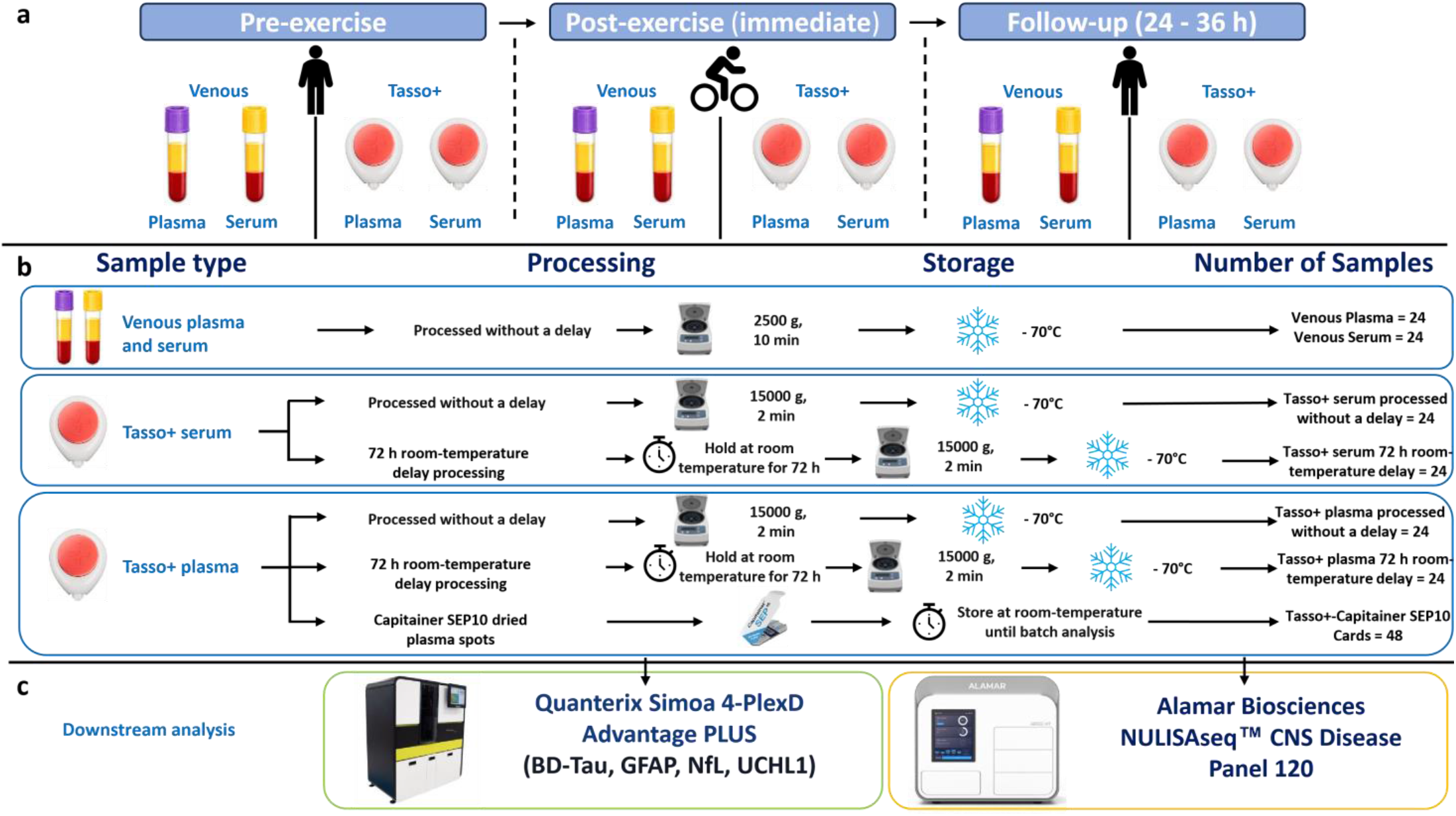
Study design and sample processing workflow. a, Blood sampling was performed in eight healthy adults at three timepoints: pre-exercise, immediately post-exercise and 24-36 h after exercise. At each timepoint, venous plasma, venous serum, Tasso+ plasma and Tasso+ serum were collected. b, Venous plasma and serum were processed without delay, centrifuged at 2,500g for 10 min and stored at -70 °C. Tasso+ plasma and serum were either processed without delay (0 h) or held at room temperature for 72 h before centrifugation at 15,000g for 2 min, after which samples were stored at -70 °C. A subset of Tasso+ plasma samples was used to prepare Capitainer SEP10 dried plasma spot cards, which were stored at room temperature until batch analysis. Sample numbers for each collection and processing condition are summarised. c, Downstream analyses were performed using the Alamar Biosciences NULISAseq CNS Disease Panel 120 and the Quanterix Simoa 4-PlexD Advantage PLUS assay for BD-Tau, GFAP, NfL and UCHL1.

### Exercise protocol

Participants completed a structured exercise session on a Wattbike cycle ergometer (Wattbike Ltd., UK). The protocol began with a 5 min self-paced warm-up. During this period, participants selected magnetic and air brake settings that were sustainable for maximal effort^15^. Heart rate was measured continuously using the same wrist-based heart monitor (Garmin Swim™ 2, Garmin Ltd., Switzerland). The warm-up was followed by a 3 min maximal effort test to determine peak power output and peak heart rate. Participants were instructed to sustain the highest achievable power over the full 3 min duration. Peak power and peak heart rate were recorded, and estimated VO_2_max was derived from the Wattbike performance algorithm that has been shown to correlate with laboratory-measured VO_2_max^15^.

Following the maximal test, participants rested until heart rate returned to baseline before completing a 25 min interval session comprising multiple high-intensity efforts separated by active recovery periods, with power targets based on performance on the previous 3 min maximal effort test.

### Blood collection

Venous and Tasso+ capillary blood samples were collected at three timepoints: pre-exercise, immediately post-exercise, and follow-up 24-to-36 h post-exercise.

### Venous blood collection and processing

Venous EDTA plasma and serum samples were collected by venepuncture. Serum samples were allowed to clot for 30 min before centrifugation at 2,500 g for 10 min. Plasma samples were centrifuged immediately at 2,500 g for 10 min. Plasma and serum were aliquoted (1.5 ml) and stored at −70 °C until batch analysis. Before analysis, venous blood aliquots were thawed at room temperature, briefly vortexed, and centrifuged at 10,000 g at 21 °C for 5 min prior to plating.

### Tasso+ collection and processing

Tasso+ whole blood samples of approximately 500 µl were collected by trained personnel according to the manufacturer’s instructions. Capillary whole blood was collected into BD SST serum separator tubes (cat no. 365968, BD, Franklin Lakes, New Jersey,) and BD K2EDTA tubes for plasma (cat no. 365975, BD, Franklin Lakes, New Jersey). Immediately after collection and prior to centrifugation, the contents of each tube were divided into two equal aliquots: one designated for processing without a delay (0 h) and the other for processing after a 72-hour room-temperature delay (72 h). Serum aliquots were allowed to clot for 30 min prior to centrifugation. Samples were centrifuged at 15,000 g for 2 min at 21 °C to separate plasma or serum, aliquoted, and stored at −70 °C until batch analysis. Before analysis, Tasso+ plasma and serum aliquots were thawed at room temperature, briefly vortexed, and centrifuged at 10,000 g at 21 °C for 5 min prior to plating.

### Capitainer SEP10 cards collection and processing

Capitainer SEP10 cards (cat no. 110-0050, Capitainer AB, Solna, Sweden) were prepared by pipetting 70 µL of whole blood from the EDTA Tasso+ collection tube prior to centrifugation (Capitainer-Tasso+). Once the blood reached the fill stop line, the card was held at a 90° angle with the sample inlet facing upwards for 2 seconds, then returned to a flat position on the bench. Following completion of plasma separation, indicated by a blue colour change in the observation window, the card was closed and placed on a Capitainer drying rack with the inlet facing downwards. Cards were allowed to dry overnight at room temperature, returned to their original packaging, and stored at room temperature until batch analysis (31.5 ± 23.0 days).

The SIMOA dried plasma spot (DPS) extraction kit (cat no. 105909, Quanterix, Billerica, MA) was used for analysis; 170 µL of DPS extraction buffer was added to each well of a precipitation plate containing two Capitainer SEP10 DPS discs, which was stacked on top of a 96-well collection plate. Plates were incubated at 37 °C for 30 min with shaking at 550 rpm, followed immediately by centrifugation at 2,500g for 15 min at room temperature. A volume of 130 µL of eluate was transferred to a fresh sample plate for analysis, which was performed according to the manufacturer’s protocol.

For NULISAseq analysis, 120 µL of NULISA DPS/DBS extraction buffer was added to each well containing two Capitainer-Tasso+ DPS discs in a precipitation plate stacked on top of a 96-well collection plate. Plates were incubated at room temperature for 1 h with shaking at 1,000 rpm, followed by centrifugation at 2,500 g for 15 min at room temperature. A volume of 55 µL of eluate was transferred to a fresh sample plate for analysis. NULISAseq assays were performed using an adapted version of the manufacturer’s protocol, optimized for Capitainer SEP10 cards.

### Immunoassay protocol and quality control

Quantification of biomarkers was performed using the Quanterix SIMOA 4plexD+ (catalog number: 104822, Quanterix, Billerica, MA), following the manufacturer’s instructions. Venous plasma and serum samples and kit-provided controls were analysed using the instrument on board dilution of 1:4, with study samples run in singlicate and controls run in duplicate. Capitainer-Tasso+ eluates were analysed in singlicate using a neat (undiluted) protocol. To assess intra-plate technical variability, 8 replicates of pooled venous plasma were included on each assay plate. Intra-plate precision, expressed as coefficient of variation (%CV), was 4.9% for brain-derived tau (BD-Tau), 5.2% for GFAP, 9.1% for NfL, and 7.8% for UCHL1.

Biomarkers were also analysed using the Alamar Biosciences NULISAseq CNS Disease panel 120 v2 (cat no. 800104, Alamar Biosciences, Fremont, CA) following the manufacturer’s instructions. Samples were processed using the *CNS_Disease_Panel_120_(v2) 55ul* protocol, with Capitainer-Tasso+ samples run neat and venous plasma and serum samples run at a 1:4 dilution using Alamar sample diluent buffer (cat no. 801352, Alamar Biosciences, Fremont, CA). The next-generation sequencing (NGS) library was sequenced on an Illumina NextSeq 2000 platform (Illumina, San Diego, CA, USA) using XLEAP-SBS P2 100-cycle chemistry (cat no. 20100987, Illumina, San Diego, CA, USA) according to a custom sequencing recipe provided by Alamar Biosciences. Inter-plate assay precision for the panel, assessed using kit-provided controls was 8.17%. All samples were run in the same experiment for both assays.

### Statistical analysis

All statistical analyses were performed in R version 4.4.3.

For each biomarker and method comparison, the number of paired observations (n), median, and interquartile range (IQR) of reference values were calculated. Method correlations were assessed using Spearman’s rank correlation coefficient (r_s_) with 95% confidence intervals estimated using non-parametric bootstrapping, two-sided p-values, and method agreement was further evaluated using Lin’s concordance correlation coefficient (CCC) reported with 95% confidence intervals. Given the small sample size, emphasis was placed on point and interval estimates and pre-specified acceptability criteria rather than null-hypothesis significance testing for method agreement.

Differences in SIMOA measurements were summarised as median percentage change relative to the relevant reference condition and reported with interquartile ranges. Concordance between sampling methods was assessed using Spearman rank correlation. Agreement was further evaluated using Bland-Altman analysis, in which difference in concentration (pg/mL) was plotted against mean concentration (pg/mL), and mean bias and limits of agreement (mean ± 1.96 SD) were estimated with 95% confidence intervals derived by nonparametric bootstrap resampling. Due to the number of paired samples being limited, the limits of agreement and their 95% bootstrap confidence intervals were expected to be wide and are reported for transparency. For NULISAseq data, differences between methods were summarised as the median log_2_ fold change (log_2_FC), defined as the difference between test and reference NPQ (NULISA protein quantification) values (ΔNPQ=test NPQ − reference NPQ), with corresponding IQR. Agreement for NULISAseq data was further visualised using two-dimensional summary plots of r_s_ versus median ΔNPQ for each protein, with predefined thresholds overlaid to indicate acceptability categories and selected key proteins annotated. Biomarker performance was classified as uncertain when ≥50% of reference samples fell below the lower limit of quantification (LLOQ) + 0.5 NPQ; otherwise, performance was classified as high (|ΔNPQ| ≤ 0.5 and r_s_ ≥ 0.75), moderate (|ΔNPQ| ≤ 0.75 and 0.50 ≤ r_s_ < 0.75), or low (|ΔNPQ| > 0.75 or r_s_ < 0.50). Although these are arbitrary cut-points, they were chosen pragmatically to support interpretation of relative bias and rank-order agreement in this exploratory analysis. Samples that did not yield a reportable measurement value were excluded from downstream analyses for NULISAseq and SIMOA markers. For NULISAseq APOE and CRP, no measurements were obtained for Capitainer-Tasso+ samples or Tasso+ plasma processed with a delay (72 h), these analytes were therefore excluded from agreement analyses.

Exercise-associated changes in NULISAseq and SIMOA markers were analysed using linear mixed-effects models with timepoint (pre-exercise, post-exercise, follow-up) as a categorical fixed effect and participant as a random intercept, fitted separately within each sample condition. Estimated marginal means were used to derive pairwise contrasts between timepoints. Effect sizes are reported as ΔNPQ for NULISAseq and as absolute differences in concentration for SIMOA, each with 95% confidence intervals. P-values were adjusted using the Benjamini-Hochberg false discovery rate (FDR), with FDR-adjusted p < 0.05 considered significant. Volcano plots were used to visualise exercise-associated changes in NULISAseq markers. SIMOA-measured venous plasma and serum UCHL1 were excluded from the exercise analysis due to insufficient paired observations.

## Results

Healthy adult participant characteristics (n=8) are summarised in **Sup Table 1**.

### Agreement between venous and Tasso+ samples processed without a delay (0 h)

To establish baseline effects associated with the Tasso+ device, Tasso+ samples processed without a delay (0 h) compared with matched venous plasma and serum reference samples. SIMOA GFAP showed strong agreement with venous samples in plasma (Spearman’s rank correlation (r_s_) = 0.95, 95% confidence interval (CI) 0.87-0.99; -1.73%) and serum (r_s_=0.98, 95% CI 0.91-1.00; 3.88%) (**Sup Table 2, *Sup Fig***. ***1***). NfL exhibited moderate agreement in plasma (r_s_=0.69, 95% CI 0.44-0.83; −7.99%) and stronger agreement in serum (r_s_=0.80, 95%CI 0.58-0.90; 4.95%), whereas BD-Tau exhibited weak rank correlation and pronounced positive bias in both plasma (r_s_=0.02, 95% CI -0.43-0.42; 1102.01%) and serum (r_s_=0.07, 95% CI -0.43-0.48; 1078.91%), indicating that concentrations measured in Tasso+ samples were substantially higher than those measured in matched venous samples. SIMOA-based UCHL1 analysis was limited by venous reference concentrations falling below the assay quantification range, leaving only 10 paired observations for stability analysis and showing low agreement with substantial bias in plasma (r_s_=0.44, 95% CI -0.31-0.92; 1254.61%) and serum (r_s_=0.56, 95% CI -0.04-0.89; 730.10%).

For NULISAseq analysis in plasma, 22.1% of proteins exhibited high agreement and 19.1% moderate agreement, with 43.5% classified as low and 15.3% as uncertain (NULISAseq abbreviations, **Sup Table 3**) (**Sup Table 4, Figure 2**). In serum, a greater proportion of proteins showed high agreement (32.8%) and moderate agreement (19.8%), whereas 32.1% were classified as low and 15.3% as uncertain. Several key pTau analytes, including BD-pTau-181, BD-pTau-217, BD-pTau-231, pTau-181, pTau-217 and pTau-231, showed pronounced positive median ΔNPQ when processed without a delay (0 h), indicating higher NPQ values in Tasso+ samples than in matched venous samples despite the absence of processing delay (ΔNPQ 1.03 to 4.79). This pattern reflects that observed for SIMOA measured BD-Tau (**Sup Table 2**), where Tasso+ measurements were consistently higher than matched venous values.

**Figure 2.**
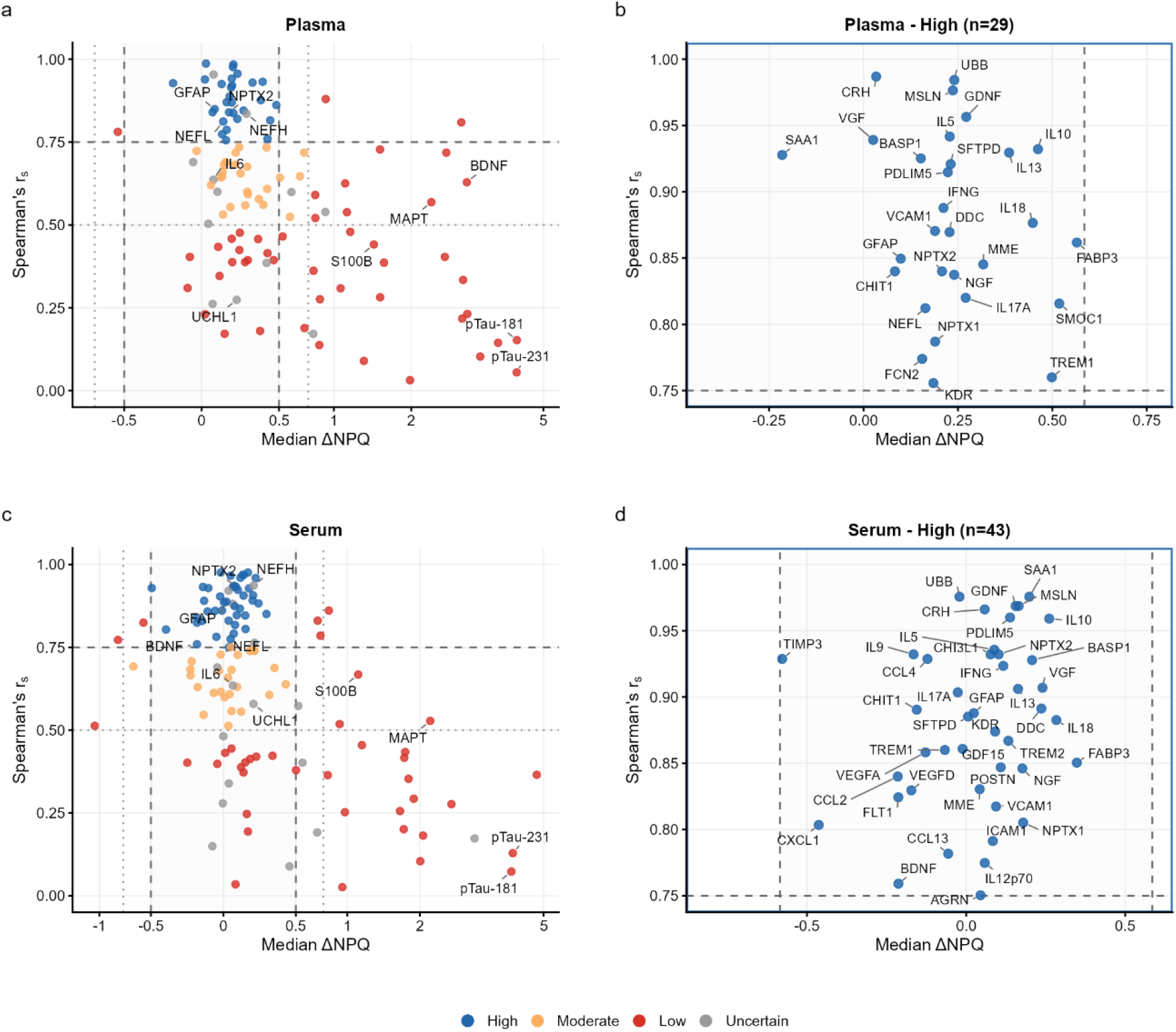
NULISAseq: venous versus Tasso+ microsampling processed without a delay (0 h). Scatter plots show agreement between venous and Tasso+ samples processed without a delay (0 h), measured by NULISAseq in plasma (a, b) and serum (c, d). Panels a and c display all quantified proteins, while panels b and d show zoomed views of proteins meeting high performance criteria. Each point represents one protein. The x-axis indicates the median difference in NPQ between methods (ΔNPQ), and the y-axis shows Spearman’s rank correlation coefficient (r_s_) across paired samples. Dashed lines denote predefined agreement thresholds (|ΔNPQ| ≤ 0.5 and r_s_ ≥ 0.75). Biomarker performance was classified as uncertain if ≥50% of reference samples were below LLOQ + 0.5 NPQ; otherwise as high (|ΔNPQ| ≤ 0.5 and r_s_ ≥ 0.75), moderate (|ΔNPQ| ≤ 0.75 and 0.50 ≤ r_s_ < 0.75), or low (|ΔNPQ| > 0.75 or r_s_ < 0.50).

### Agreement between venous and Tasso+ samples processed with a 72-hour delay

Evaluation of Tasso+ samples processed with a delay (72 h) showed robust stability for SIMOA-measured GFAP in both plasma (r_s_=0.94, 95% CI 0.84-0.98; 7.21%) and serum (r_s_=0.97, 95% CI 0.89-0.99; 8.78%), with NfL also demonstrating stability in plasma (r_s_=0.76, 95% CI 0.55-0.88; 9.58%) and serum (r_s_=0.87, 95% CI 0.72-0.95; 6.87%) (**Sup Table 5, Figure 3**). Broad-scale NULISAseq profiling further showed that approximately 35% of proteins achieved high or moderate agreement across plasma and serum; in plasma, 20.2% of proteins were classified as high and 14.7% as moderate agreement, while in serum 19.8% were classified as high and 15.3% as moderate agreement (**Figure 4, Sup Table 6**).

**Figure 3.**
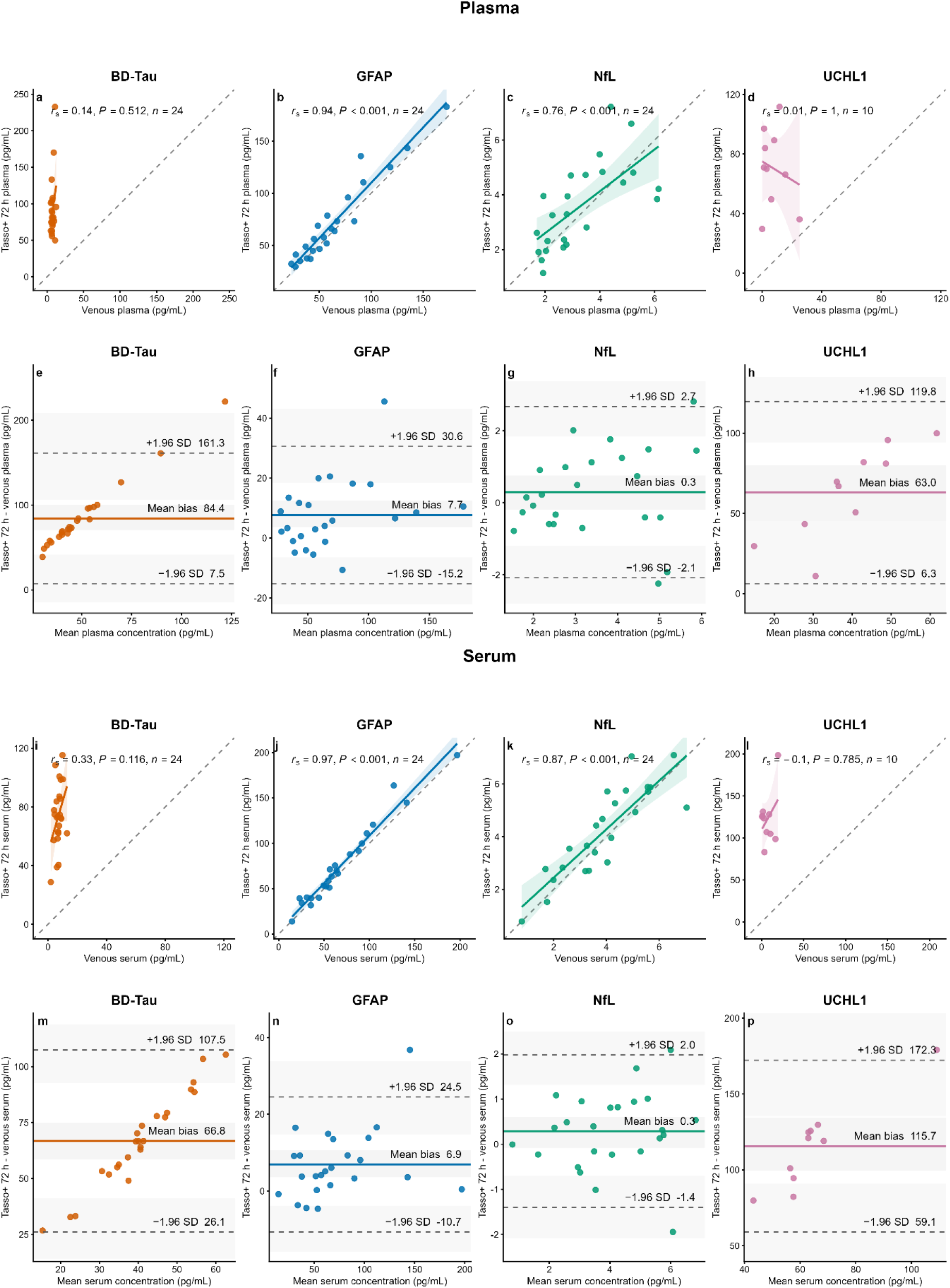
SIMOA: Venous versus Tasso+ plasma and serum processed with a delay (72 h). Plasma: (a-d) Spearman correlation plots comparing venous and Tasso+ 72 h plasma concentrations for BD-Tau, GFAP, NfL and UCHL1. (e-h) Bland-Altman plots showing the difference in concentration between Tasso+ 72 h and venous plasma samples, plotted against their mean concentration. Serum: (i-l) Spearman correlation plots comparing venous and Tasso+ 72 h serum concentrations for the same analytes. (m-p) Bland-Altman plots showing the difference in concentration between Tasso+ 72 h and venous serum samples, plotted against their mean concentration. For correlation plots, dashed lines indicate the line of identity and solid lines show linear regression fits with 95% confidence intervals; Spearman’s r_s_, two-sided P value and n are shown. For Bland-Altman plots, solid lines indicate mean bias, dashed lines indicate limits of agreement (mean ± 1.96 SD), and shaded bands represent 95% bootstrap confidence intervals. Tasso+ 72 h denotes samples processed after a 72 h room-temperature delay.

**Figure 4.**
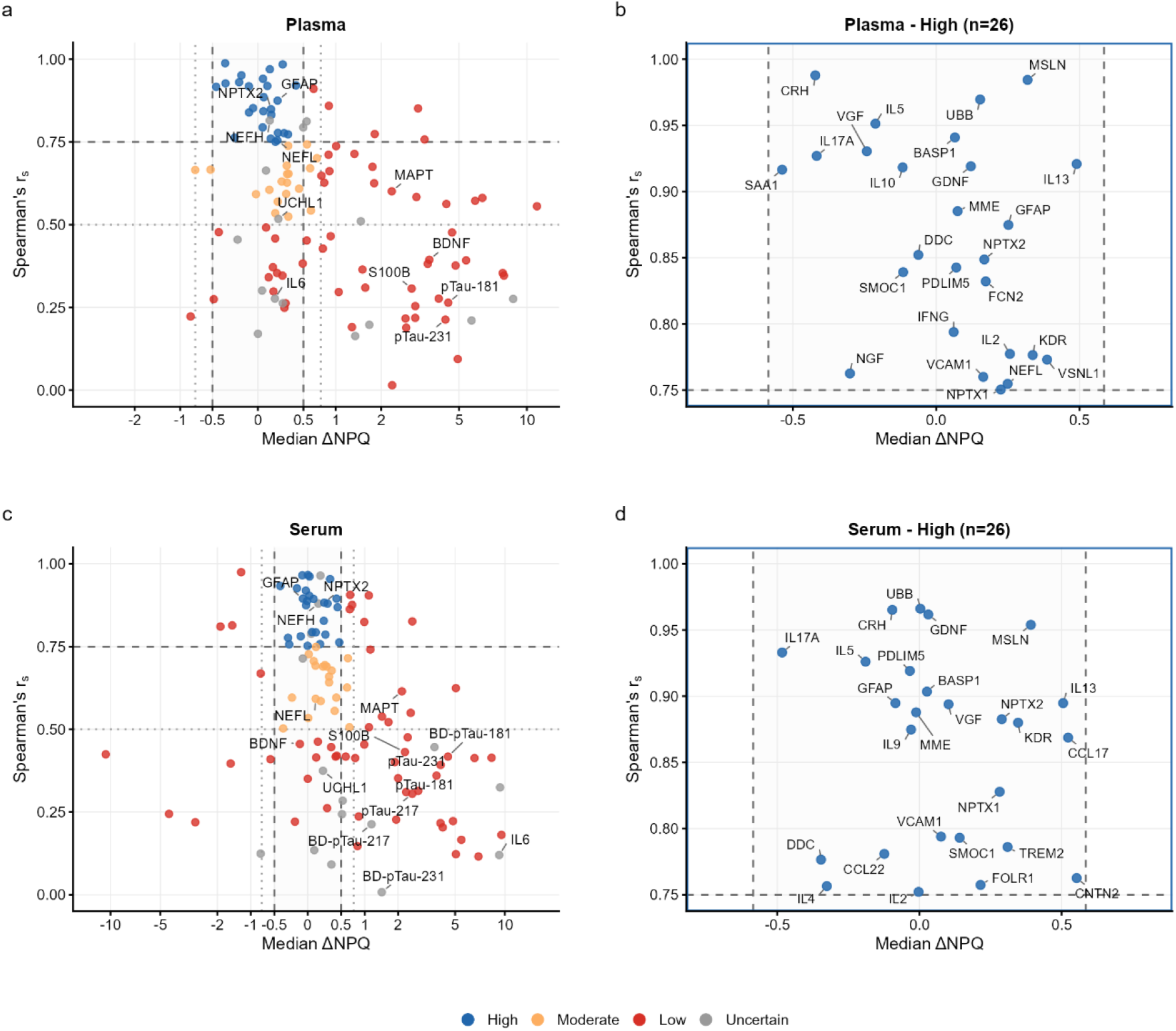
NULISAseq: venous versus Tasso+ microsampling processed with a delay (72 h). Scatter plots show agreement between venous and Tasso+ samples processed with a delay (72 h), measured by NULISAseq in plasma (a, b) and serum (c, d). Panels a and c display all quantified proteins, while panels b and d show zoomed views of proteins meeting high performance criteria. Each point represents one protein. The x-axis indicates the median difference in NPQ between methods (ΔNPQ), and the y-axis shows Spearman’s rank correlation coefficient (r_s_) across paired samples. Dashed lines denote predefined agreement thresholds (|ΔNPQ| ≤ 0.5 and r? ≥ 0.75). Biomarker performance was classified as uncertain if ≥50% of reference samples were below LLOQ + 0.5 NPQ; otherwise as high, moderate, or low according to the predefined bias and correlation criteria.

Among NULISAseq analytes assessed in Tasso+ samples processed with a delay (72 h), GFAP and NPTX2 showed high agreement with matched venous samples in both plasma and serum. High agreement was observed for serum GFAP (r_s_=0.89, 95% CI 0.72-0.96; ΔNPQ = -0.06), NPTX2 (r_s_=0.88, 95% CI 0.73-0.94; ΔNPQ = 0.22), and plasma GFAP (r_s_=0.87, 95% CI 0.64, 0.97; ΔNPQ=0.19), NPTX2 (r_s_=0.85, 95% CI 0.68-0.93; ΔNPQ = 0.12), and NfL (r_s_ = 0.75, 95% CI 0.51-0.89; ΔNPQ = 0.19) meeting criteria for high agreement. While NfL reached high agreement in plasma, serum NfL displayed clear platform-dependent variance, maintaining stability by SIMOA but showing reduced agreement on NULISAseq (r_s_=0.59, 95% CI 0.28-0.79; ΔNPQ=0.10). Together, these findings suggest that differences between assay platforms may influence agreement independently of whether the analyte performs consistently in plasma or serum.

### Effect of a 72-hour processing delay on Tasso+ measurements

Comparison of samples processed with and without a 72-hour delay collected with the Tasso+ device revealed minimal differences for SIMOA-measured GFAP, NfL and BD-Tau in paired plasma samples and paired serum samples (**Sup Fig. 2, Sup Table 7**). GFAP and NfL showed strong concordance between processing conditions, (GFAP: plasma r_s_=0.98, 95% CI 0.92-1.00, 9.52%; serum r_s_=0.99, 95% CI 0.95-1.00, 1.67%; NfL: plasma r_s_=0.85, 95% CI 0.67-0.93, 14.76%; serum r_s_=0.87, 95% CI 0.69-0.94, -0.42%), whereas BD-Tau showed little percentage change but weak correlation (plasma r_s_=0.34, 95% CI -0.03-0.62; -0.97%; serum r_s_=0.11, 95% CI -0.32-0.54; 1.31%). In Tasso+ plasma, all key pTau ΔNPQ values ranged from -0.01 to 0.16, while in serum they ranged from -0.08 to 0.08 (**Sup Fig. 3, Sup Table 8**).

### Comparison of Capitainer-Tasso+ microsampling

Comparison of Capitainer-Tasso+ samples was assessed using Spearman’s rank correlation only, because the limited sample retained on the Capitainer SEP10 discs required elution before analysis, effectively diluting the sample to generate sufficient volume for the assays and making direct comparison of bias with Tasso+ and venous samples inappropriate. Analysis of Capitainer-Tasso+ samples using NULISAseq showed 15.5% of proteins met criteria for high agreement and 22.5% for moderate, 48% low, and 14.0% were classified as uncertain (**Sup Table 9**). Among key proteins, only NPTX2 demonstrated high agreement with venous plasma (r_s_=0.83, 95% CI 0.66-0.91). SIMOA similarly showed weak rank correlation across all measured analytes, with NfL (r_s_=-0.21, 95% CI -0.64-0.39 ; 9 pairs excluded due to inability to quantify), GFAP (r_s_=0.45, 95% CI 0.06-0.71), BD-Tau (r_s_=0.05, 95% CI -0.40 to 0.49) and UCHL1 (r_s_=0.36, 95% CI -0.40-0.81) (**Table 1**).

**Table 1.** Agreement between venous plasma and Capitainer-Tasso+ plasma measurements (SIMOA). Comparison of venous plasma and Capitainer-Tasso+ plasma marker measurements quantified using the Quanterix SIMOA 4-PlexD Advantage PLUS assay for BD-Tau, GFAP, NfL and UCHL1. Values are reported as median (IQR). Associations were assessed using Spearman’s rank correlation coefficient (**r**_**s**_) with two-sided p values.

| Marker | n (pairs) | Venous plasma | Capitainer-Tasso+ | $r_s$ | P |
| --- | --- | --- | --- | --- | --- |
|  |  | Median (IQR) | Median (IQR) |  |  |
| BD-Tau | 24 | 7.04 (1.98) | 7.62 (4.69) | 0.05 | 0.799 |
| GFAP | 24 | 55.99 (38.34) | 3.46 (4.13) | 0.45 | 0.027 |
| NfL | 15 | 2.94 (1.79) | 0.26 (1.38) | -0.21 | 0.459 |
| UCHL1 | 10 | 4.58 (9.35) | 39.14 (25.41) | 0.36 | 0.31 |

### Effects of Exercise

Exercise-associated biomarker effects were most evident in samples taken immediately post-exercise. Using SIMOA, GFAP decreased after exercise in plasma, reaching significance in Tasso+ plasma processed without a delay (0 h) (Δ -15.97 pg/mL, 95% CI -26.61 to -5.32; FDR-adjusted *P* = 0.025) and venous plasma (Δ -22.55 pg/mL, 95% CI -39.71 to -5.39; FDR-adjusted *P* = 0.041) (**Sup Table 10**). SIMOA-measured BD-Tau increased immediately after exercise in venous serum (Δ 2.11 pg/mL, 95% CI 0.54 to 3.67; FDR-adjusted *P* = 0.036) and increased further between the post-exercise and 24-to-36 h follow-up timepoints (Δ 0.76 pg/mL, 95% CI 0.42 to 1.10; FDR-adjusted *P* = 0.001). UCHL1 also increased acutely in Tasso+ plasma processed without a delay (0 h) (Δ 21.65 pg/mL, 95% CI 4.00 to 39.30; FDR-adjusted *P* = 0.040). No markers measured with SIMOA differed significantly between pre-exercise sampling and 24-36 h post-exercise following FDR correction.

NULISAseq identified broader but sample-type-specific acute exercise responses, which were most evident in venous plasma (**Figure 5, Sup Table 11**). Post-exercise increases included BDNF (ΔNPQ 1.70, 95% CI 0.90 to 2.51; FDR-adjusted *P* = 0.009) and IL6 (ΔNPQ 0.41, 95% CI 0.24 to 0.57; FDR-adjusted *P* = 0.003), while GFAP decreased in Tasso+ plasma processed without a delay (0 h) (ΔNPQ -0.39, 95% CI -0.59 to -0.20; FDR-adjusted *P* = 0.032) (Sup Table 11). Significant changes detected in other Tasso+ conditions were more limited and included increased FLT1 in Tasso+ plasma and serum processed without a delay (0 h). Most NULISAseq changes were transient, with many markers demonstrating decreases immediately post-exercise to 24-to-36 h follow-up, particularly in venous plasma, including BDNF (ΔNPQ -1.82, 95% CI -2.62 to -1.02; FDR-adjusted *P* = 0.004), IL6 (ΔNPQ -0.40, 95% CI -0.56 to - 0.24; FDR-adjusted *P* = 0.003) and TIMP3 (ΔNPQ -1.81, 95% CI -2.45 to -1.16; FDR-adjusted *P* = 0.001).

**Figure 5.**
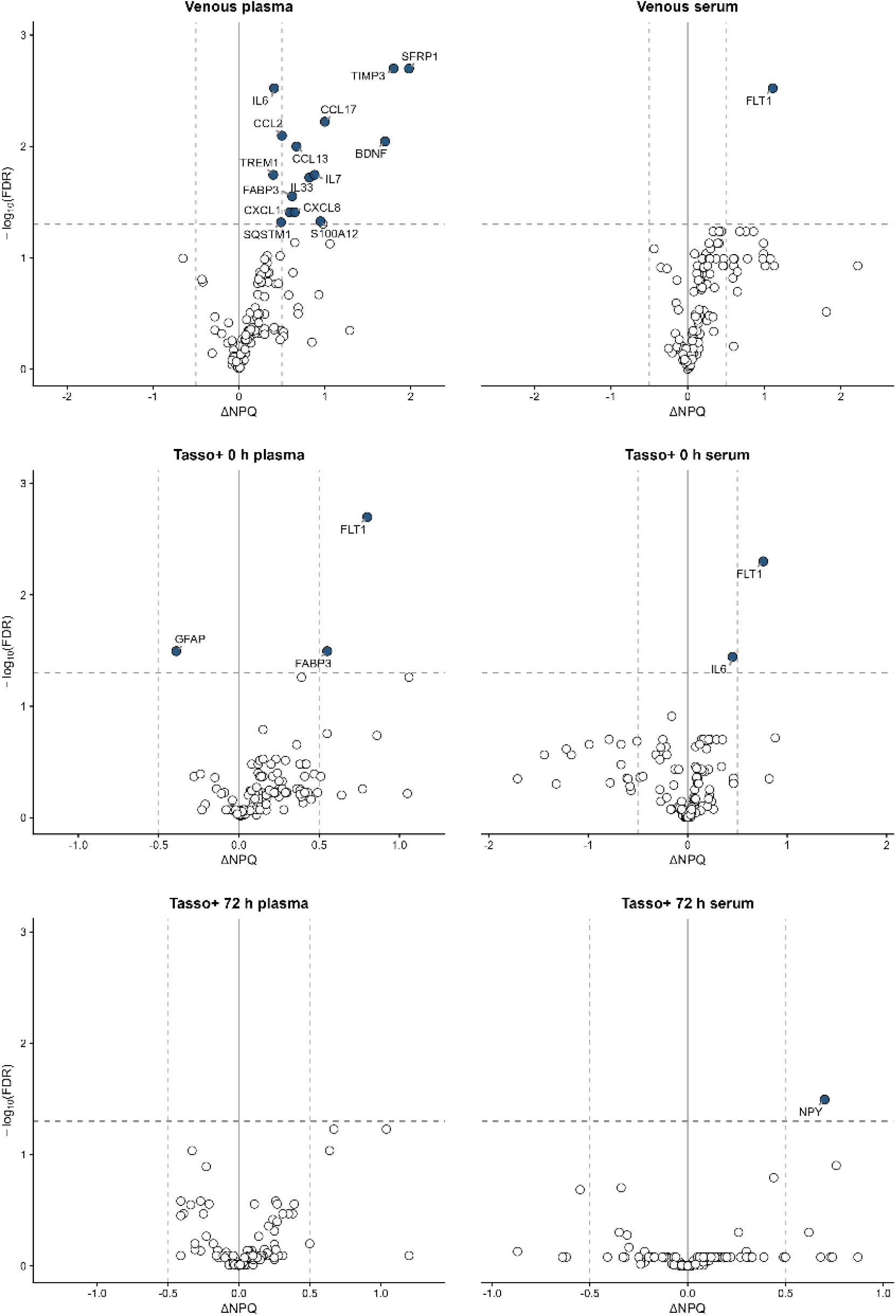
Exercise-associated changes in circulating CNS-related proteins measured by NULISAseq. Volcano plots comparing post-exercise with pre-exercise in venous plasma, venous serum, Tasso+ plasma processed without a delay (0 h), Tasso+ serum processed without a delay (0 h), Tasso+ plasma processed with a delay (72 h), and Tasso+ serum processed with a delay (72 h). Proteins are plotted by ΔNPQ, calculated as post-exercise minus baseline, and -log10 false discovery rate (FDR)-adjusted P value. Horizontal dashed lines mark the FDR significance threshold, and vertical dashed lines mark ΔNPQ values of ±0.5. Labelled proteins met the FDR significance threshold. Positive ΔNPQ values indicate increased abundance after exercise, whereas negative values indicate decreased abundance.

Consistent with return towards pre-exercise, only one NULISAseq marker remained significantly different at 24-to-36 h post-exercise compared to pre-exercise after FDR correction: IL7 in Tasso+ serum processed without a delay (0 h) (ΔNPQ -0.61, 95% CI -0.83 to -0.39; FDR-adjusted *P* = 0.005).

## Discussion

We show that the Tasso+ device supports ultrasensitive CNS biomarker measurement in matched plasma and serum samples processed without a delay (0 h) or with a 72 h room-temperature delay (72 h). Using SIMOA, capillary GFAP and NfL retained strong concordance with matched venous reference samples across both plasma and serum under both processing conditions. As expected, NULISAseq showed more variable analyte-level agreement, with high or moderate agreement observed for 41.2% of plasma proteins (high: 22.1%, moderate: 19.1%) and 52.6% of serum proteins (high: 32.8%, moderate: 19.8%) when processed without a delay (0 h), and 34.9% of plasma proteins (high: 20.2%, moderate: 14.7%) and 35.1% of serum proteins (high: 19.8%, moderate: 15.3%) when processed with a delay (72 h). This study provides the first combined evaluation of Tasso+ plasma and serum when processed without a delay (0 h) and with a delay (72 h) using the NULISAseq CNS Disease Panel 120 and 4-PlexD+ SIMOA, with suitability depending on the proteins of interest.

Our finding that SIMOA-measured NfL and GFAP remain robust when Tasso+ capillary samples are processed with a delay (72 h) is consistent with prior finger-prick microsampling studies showing strong agreement after 3 days ^3^. Prior venous stability studies have also shown that pTau-181 and pTau-217 remain stable over 72 h ^10,11^. However, those studies only assessed change in concentration, whereas we assessed both change over time and correlation agreement with matched venous samples. Consistent with limited time-dependent degradation, comparisons of Tasso+ samples processed without a delay (0 h) and with a delay (72 h) showed only small ΔNPQ for pTau analytes. However, correlation between Tasso+ and venous samples was poor for several pTau analytes under both processing conditions. This suggests that the lack of agreement is unlikely to be explained by processing delay alone. Instead, it may reflect the low pTau concentrations and narrow biological range in this healthy cohort, which limit the ability to detect correlation between sample types. Correlation agreement may therefore be clearer in populations with higher pTau concentrations ^4^. However, it is important to note that this study does not refute that proteins showing low or uncertain agreement may be unsuitable for Tasso+, but further studies in disease-relevant populations are required to fully evaluate their performance.

We examined exercise-associated changes across the biomarker panel to assess whether recent high-intensity exercise could confound interpretation in sport-related sampling contexts. Most biomarkers did not show statistically significant changes across timepoints after FDR adjustment. Previous studies suggest that exercise-related GFAP changes are acute and short-lived, with reported immediate post-exercise decreases returning to baseline within 45-mins to approximately 1 h ^9,12^. In our study, GFAP decreased immediately post-exercise in venous plasma and Tasso+ plasma processed without a delay (0 h), returning toward pre-exercise by the 24-to-36 h follow-up. Importantly, this direction of change contrasts with sport-related concussion studies, where GFAP is typically increased after injury, including acutely and at 24-48 hours post-injury ^13^. Exercise-related GFAP reductions are therefore unlikely to produce false-positive injury signals. BDNF increases observed immediately post-exercise returned to pre-exercise by the 24-to-36 h follow-up, consistent with previous evidence that acute exercise increases circulating BDNF ^14^. Subtle increases in BD-Tau were also observed, but this has not been reported previously and requires validation in larger studies, particularly as the finding was not statistically significant across all sampling conditions.

The study population consisted of young, healthy individuals, while the biomarkers examined are typically elevated in neurodegenerative and neuroinflammatory disorders, meaning the study may be underpowered in terms of both cohort composition and expected biomarker concentrations. This was particularly relevant for the Capitainer-Tasso+ comparison, in which many proteins were not detectable, and stronger correlations may emerge in populations with higher biomarker concentrations.^4^ In addition, measurements were restricted to the NULISAseq and SIMOA platforms and biomarker performance may vary across analytical technologies ^11^. Future work should include larger validation cohorts, including in individuals with established neurological conditions, and assess a broader range of assays.

Overall, this study provides evidence that several key markers of brain health can be reliably assessed using Tasso+ even when processed with a delay (72 h). This is particularly relevant for field-based research in which conventional venepuncture is logistically challenging, such as studies examining the effects of sporting-related head impacts, as well as studies with participant-led remote sampling.

## Data Availability

All data produced in the present study are available upon reasonable request to the authors

## Sup. Figures and tables

**Sup Fig. 1. SIMOA: Venous versus Tasso+ plasma and serum processed without a delay (0 h)**. Plasma: (a-d) Spearman correlation plots comparing venous plasma with Tasso+ plasma processed without a delay (0 h) for BD-Tau, GFAP, NfL and UCHL1. (e-h) Bland-Altman plots showing the difference in concentration between Tasso+ plasma processed without a delay (0 h) and venous plasma samples, plotted against their mean concentration. Serum: (i-l) Spearman correlation plots comparing venous serum with Tasso+ serum processed without a delay (0 h) for the same analytes. (m-p) Bland-Altman plots showing the difference in concentration between Tasso+ serum processed without a delay (0 h) and venous serum samples, plotted against their mean concentration. For correlation plots, dashed lines indicate the line of identity and solid lines show linear regression fits with 95% confidence intervals; Spearman’s rank correlation coefficient (r_s_), two-sided P value and sample size (n) are shown. For Bland-Altman plots, solid lines indicate mean bias, dashed lines indicate limits of agreement (mean ± 1.96 SD), and shaded bands represent 95% bootstrap confidence intervals.

**Sup Fig. 2. SIMOA: Tasso+ plasma and serum processed without a delay (0 h) versus with a delay (72 h)**. Plasma: (a-d) Spearman correlation plots comparing Tasso+ plasma concentrations processed without a delay (0 h) and with a delay (72 h) for BD-Tau, GFAP, NfL and UCHL1. (e-h) Bland-Altman plots showing the difference in concentration between Tasso+ plasma samples processed with a delay (72 h) and without a delay (0 h), plotted against their mean concentration. Serum: (i-l) Spearman correlation plots comparing Tasso+ serum concentrations processed without a delay (0 h) and with a delay (72 h) for the same analytes. (m-p) Bland-Altman plots showing the difference in concentration between Tasso+ serum samples processed with a delay (72 h) and without a delay (0 h), plotted against their mean concentration. For correlation plots, dashed lines indicate the line of identity and solid lines show linear regression fits with 95% confidence intervals; Spearman’s rank correlation coefficient (r_s_), two-sided P value and sample size (n) are shown. For Bland-Altman plots, solid lines indicate mean bias, dashed lines indicate limits of agreement (mean ± 1.96 SD), and shaded bands represent 95% bootstrap confidence intervals.

**Sup Fig. 3. NULISAseq: Tasso+ plasma and serum processed without a delay (0 h) versus with a delay (72 h)**. Scatter plots show agreement between Tasso+ samples measured by NULISAseq in plasma (**a, b**) and serum (**c, d**) processed without a delay (0 h) and with a delay (72 h); panels **a** and **c** display all quantified proteins and panels **b** and **d** show zoomed views of proteins meeting High performance criteria. Each point represents one protein. The x-axis indicates the median ΔNPQ between processing conditions and the y-axis shows Spearman’s r_s_ across paired samples. Dashed lines denote predefined agreement thresholds (|ΔNPQ| ≤ 0.5 and r_s_ ≥ 0.75). Biomarker performance was classified as uncertain if ≥50% of reference samples were below LLOQ + 0.5 NPQ; otherwise as high, moderate or low according to the predefined bias and correlation criteria.

**Sup Table 1. Participant characteristics**. Participant demographic and clinical characteristics are summarised for the study cohort. Categorical variables are reported as counts. Continuous variables are reported as median (interquartile range, IQR).

**Sup Table 2. Agreement between venous and Tasso+ plasma and serum measurements processed without a delay (0 h) (SIMOA)**. Comparison of venous and Tasso+ plasma and serum marker concentrations measured using the SIMOA 4-Plex assay, with Tasso+ samples processed without a delay (0 h). Values are reported as median (IQR). Percentage change relative to venous samples is shown. Agreement was assessed using Spearman’s rank correlation coefficient (r_s_) with two-sided P values and Lin’s concordance correlation coefficient (CCC) with 95% confidence intervals. The number of paired samples is reported for each marker. SIMOA markers include brain-derived tau (BD-Tau), glial fibrillary acidic protein (GFAP), neurofilament light chain (NfL), and ubiquitin C-terminal hydrolase L1 (UCHL1), reported in pg/mL.

**Sup Table 3. Protein abbreviations used in the Alamar Biosciences NULISAseq CNS Disease Panel**. The table lists abbreviated protein names, their corresponding full protein names, and the biological category.

**Sup Table 4. Agreement between venous and Tasso+ plasma and serum measurements processed without a delay (0 h) (NULISAseq)**. Comparison of venous and Tasso+ plasma and serum marker measurements quantified using the Alamar NULISAseq CNS Disease Panel, with Tasso+ samples processed without a delay (0 h). Differences are reported as ΔNPQ (IQR). Associations were assessed using Spearman’s rank correlation coefficient (r_s_) with two-sided P values, and agreement was evaluated using Lin’s concordance correlation coefficient (CCC) with 95% confidence intervals. Agreement classification is reported as high, moderate, low, or uncertain.

**Sup Table 5. Agreement between venous and Tasso+ plasma and serum measurements processed with a delay (72 h) (SIMOA)**. Comparison of venous and Tasso+ plasma and serum marker concentrations measured using the SIMOA 4-Plex assay, with Tasso+ samples processed with a delay (72 h). Values are reported as median (IQR), with percentage change relative to venous samples. Agreement was assessed using Spearman’s rank correlation coefficient (r_s_) with two-sided P values and Lin’s concordance correlation coefficient (CCC) with 95% confidence intervals. The number of paired samples is reported for each marker. SIMOA markers are reported in pg/mL.

**Sup Table 6. Agreement between venous and Tasso+ plasma and serum measurements processed with a delay (72 h) (NULISAseq)**. Comparison of venous and Tasso+ plasma and serum marker measurements quantified using the Alamar NULISAseq CNS Disease Panel, with Tasso+ samples processed with a delay (72 h). Differences are reported as ΔNPQ (IQR). Associations were assessed using Spearman’s rank correlation coefficient (r_s_) with two-sided P values, and agreement was evaluated using Lin’s concordance correlation coefficient (CCC) with 95% confidence intervals. Agreement classification is reported as high, moderate, low, or uncertain.

**Sup Table 7. Agreement between Tasso+ plasma and serum measurements processed without a delay (0 h) and with a delay (72 h) (SIMOA)**. Comparison of marker concentrations between Tasso+ plasma and serum samples measured using the SIMOA 4-Plex assay and processed without a delay (0 h) or with a delay (72 h). Values are reported as median (IQR), with percentage change relative to samples processed without a delay (0 h). Agreement was assessed using Spearman’s rank correlation coefficient (r_s_) with two-sided P values and Lin’s concordance correlation coefficient (CCC) with 95% confidence intervals. The number of paired samples is reported for each marker. SIMOA markers are reported in pg/mL.

**Sup Table 8. Agreement between Tasso+ plasma and serum measurements processed without a delay (0 h) and with a delay (72 h) (NULISAseq)**. Comparison of marker measurements between Tasso+ plasma and serum samples quantified using the Alamar NULISAseq CNS Disease Panel and processed without a delay (0 h) or with a delay (72 h). Differences are reported as ΔNPQ (IQR). Associations were assessed using Spearman’s rank correlation coefficient (r_s_) with two-sided P values, and agreement was evaluated using Lin’s concordance correlation coefficient (CCC) with 95% confidence intervals. Agreement classification is reported as high, moderate, low, or uncertain.

**Sup Table 9. Agreement between Capitainer-Tasso+ and venous plasma measurements (NULISAseq)**. Comparison of marker measurements between Capitainer-Tasso+ and venous plasma samples quantified using the Alamar NULISAseq CNS Disease Panel. Differences are reported as ΔNPQ (IQR). Associations were assessed using Spearman’s rank correlation coefficient (r_s_) with two-sided P values. Agreement classification is reported as high, moderate, low, or uncertain.

**Sup Table 10. Exercise-related changes in markers (SIMOA)**. Exercise-related changes in marker concentrations were assessed using linear mixed-effects models comparing pre-exercise, post-exercise, and follow-up time points. Results are reported as changes in concentration (Δ concentration) with 95% confidence intervals, unadjusted P values, and false discovery rate-adjusted P values. SIMOA markers include BD-Tau, GFAP, NfL, and UCHL1, reported in pg/mL.

**Sup Table 11. Exercise-related changes in markers (NULISAseq)**. Exercise-related changes in marker expression were assessed using linear mixed-effects models comparing pre-exercise, post-exercise, and follow-up time points. Results are reported as ΔNPQ with 95% confidence intervals, unadjusted P values, and false discovery rate-adjusted (FDR) P values.

## Conflicting Interests

MC is employed by PREM Rugby, SK and KS are employed by the Rugby Football Union, the national governing body for rugby union in England. HZ has served at scientific advisory boards and/or as a consultant for Abbvie, Acumen, Alamar, Alector, Alzinova, ALZpath, Amylyx, Annexon, Apellis, Artery Therapeutics, AZTherapies, Cognito Therapeutics, CogRx, Denali, Eisai, Enigma, LabCorp, Merck Sharp & Dohme, Merry Life, Nervgen, Novo Nordisk, Optoceutics, Passage Bio, Pinteon Therapeutics, Prothena, Quanterix, Red Abbey Labs, reMYND, Roche, Samumed, ScandiBio Therapeutics AB, Siemens Healthineers, Triplet Therapeutics, and Wave, has given lectures sponsored by Alzecure, BioArctic, Biogen, Cellectricon, Fujirebio, LabCorp, Lilly, Novo Nordisk, Oy Medix Biochemica AB, Roche, and WebMD, is a co-founder of Brain Biomarker Solutions in Gothenburg AB (BBS), which is a part of the GU Ventures Incubator Program, and is a shareholder of CERimmune Therapeutics (outside submitted work). D.J.S. has received support for blood biomarker analysis by Quanterix and Alamar. He has given a lecture with travel paid for by Alamar. He participates in an expert panel advising the English Rugby Football Union on concussion and related issues, for which he has received an honorarium from the Rugby Football Union for his participation, which has been used for research purposes. He provides expert witness reports in medical injury legal work. He has received payment for providing independent clinical care for professional players following head injuries. He has received complementary tickets to international rugby matches paid for by the Rugby Football Union.

## Sources of Funding

UK-TBI REpository and data PORTal Enabling discoveRy (TBI-REPORTER; Grant Number: MR/Y008502/1) jointly funded by the Medical Research Council (MRC), the National Institute for Health and Care Research (NIHR), the Ministry of Defence Alzheimer’s Research UK.

Traumatic Brain Injury Thresholds Study (TBI-TS) funded by the Medical Research Council (MRC) (UKRI2548).

The National Institute for Health and Care Research University College London Hospitals Biomedical Research Centre, and the UK Dementia Research Institute at UCL (UKDRI-1003).

TDP has been supported by a NIHR Clinical Lectureship, the Medical Research Council (MRC) (TBI-TS - UKRI2548), and by a UK Dementia Trials Network fellowship.

## Notes

### Author Declarations

Ethical approval was granted by the Camberwell and St Giles Research Ethics Committee, reference 17/LO/2066.

### Summary of Updates

Figures 2 and 4, and Supplementary Figure 3, have been revised.

