## Supplementary Figures 1-3 for "Analytical Validation of Minimally Invasive Capillary Blood Microsampling using Tasso+ for Multiplexed Neurological Biomarkers"

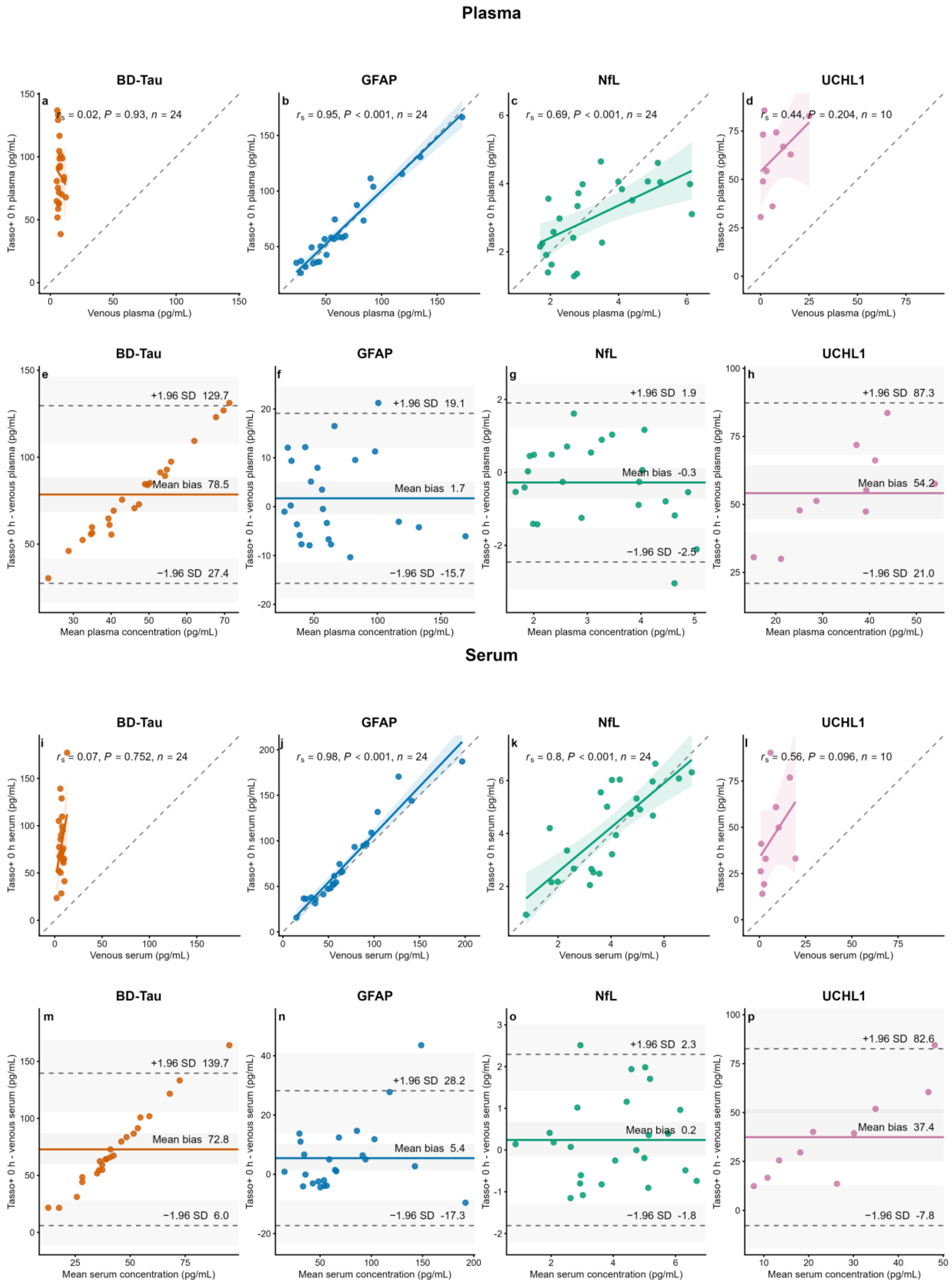


***Sup Fig. 1. SIMOA: Venous versus Tasso+ plasma and serum processed without a delay (0 h).*** *Plasma: (a-d) Spearman correlation plots comparing venous plasma with Tasso+ plasma processed without a delay (0 h) for BD-Tau, GFAP, NfL and UCHL1. (e-h) Bland-Altman plots showing the difference in concentration between Tasso+ plasma processed without a delay (0 h) and venous plasma samples,*

*plotted against their mean concentration. Serum: (i-l) Spearman correlation plots comparing venous serum with Tasso+ serum processed without a delay (0 h) for the same analytes. (m-p) Bland-Altman plots showing the difference in concentration between Tasso+ serum processed without a delay (0 h) and venous serum samples, plotted against their mean concentration. For correlation plots, dashed lines indicate the line of identity and solid lines show linear regression fits with 95% confidence intervals; Spearman's rank correlation coefficient ($r_s$), two-sided P value and sample size (n) are shown. For Bland-Altman plots, solid lines indicate mean bias, dashed lines indicate limits of agreement (mean ± 1.96 SD), and shaded bands represent 95% bootstrap confidence intervals.*

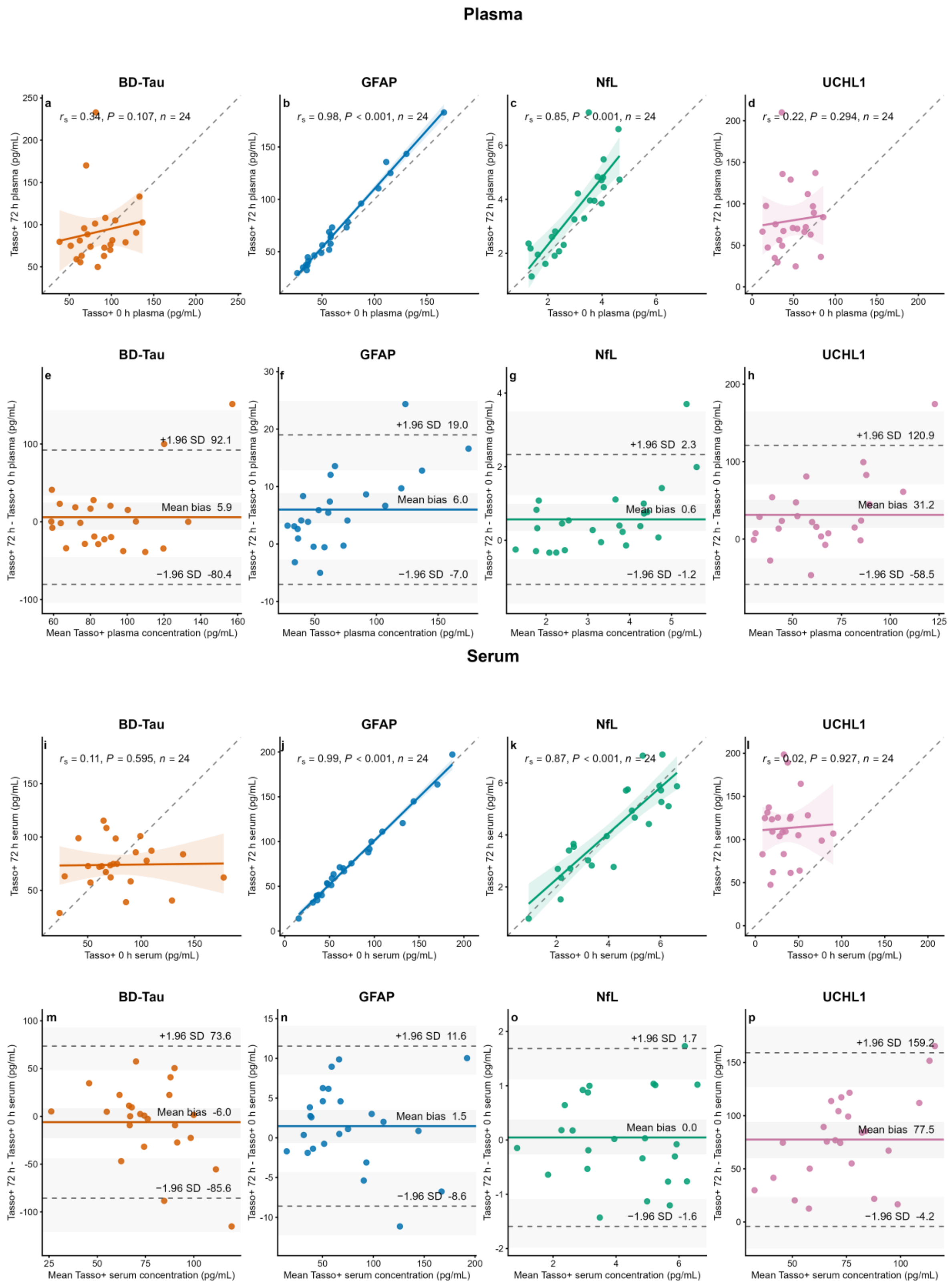


***Sup Fig. 2. SIMOA: Tasso+ plasma and serum processed without a delay (0 h) versus with a delay (72 h).*** *Plasma: (a-d) Spearman correlation plots comparing Tasso+ plasma concentrations processed without a delay (0 h) and with a delay (72 h) for BD-Tau, GFAP, NfL and UCHL1. (e-h) Bland-Altman plots showing the difference in concentration between Tasso+ plasma samples processed with a delay (72 h) and without a delay (0 h), plotted against their mean concentration. Serum: (i-l) Spearman*

*correlation plots comparing Tasso+ serum concentrations processed without a delay (0 h) and with a delay (72 h) for the same analytes. (m-p) Bland-Altman plots showing the difference in concentration between Tasso+ serum samples processed with a delay (72 h) and without a delay (0 h), plotted against their mean concentration. For correlation plots, dashed lines indicate the line of identity and solid lines show linear regression fits with 95% confidence intervals; Spearman's rank correlation coefficient ($r_s$), two-sided P value and sample size (n) are shown. For Bland-Altman plots, solid lines indicate mean bias, dashed lines indicate limits of agreement (mean ± 1.96 SD), and shaded bands represent 95% bootstrap confidence intervals.*

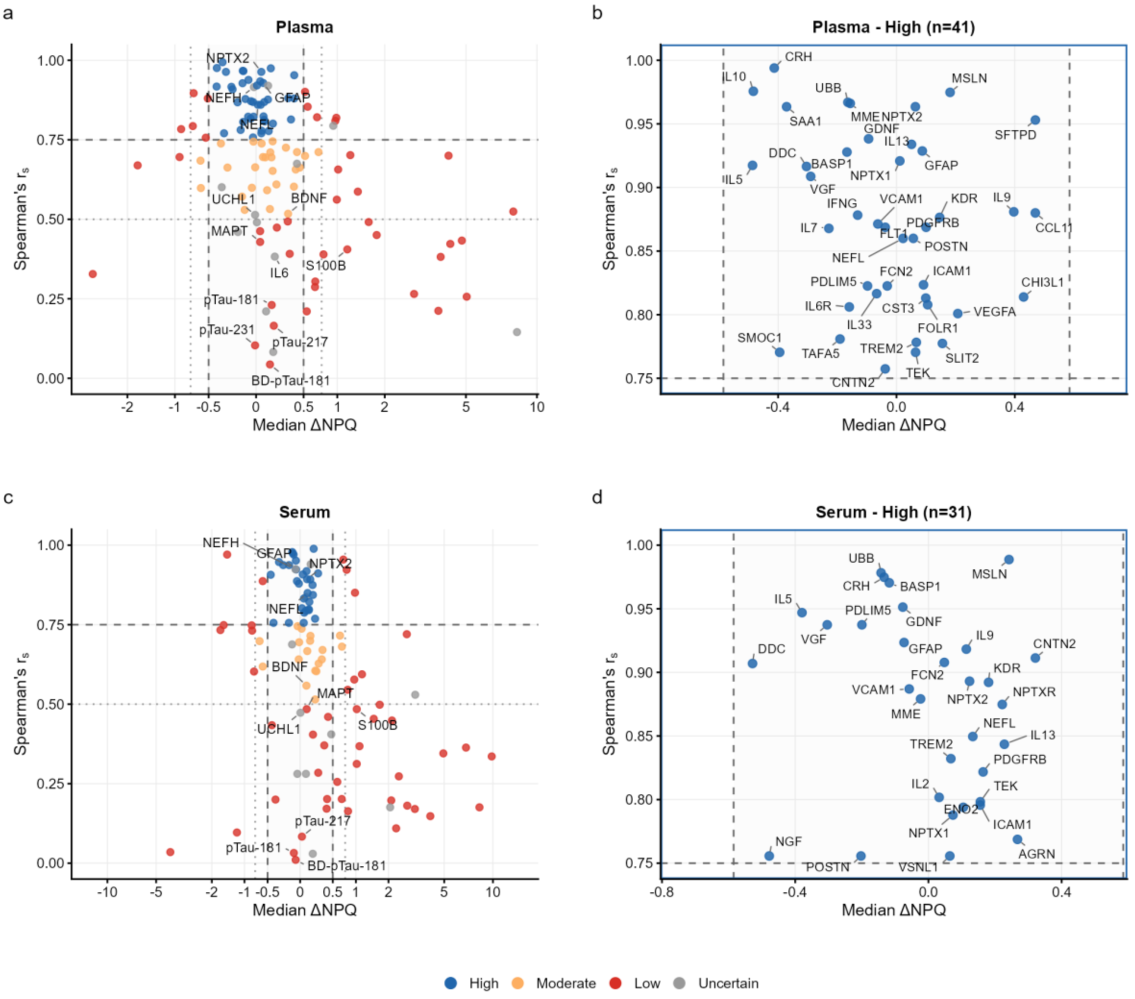


***Sup Fig. 3. NULISAseq: Tasso+ plasma and serum processed without a delay (0 h) versus with a delay (72 h).*** *Scatter plots show agreement between Tasso+ samples measured by NULISAseq in plasma (**a, b**) and serum (**c, d**) processed without a delay (0 h) and with a delay (72 h); panels **a** and **c** display all quantified proteins and panels **b** and **d** show zoomed views of proteins meeting High performance criteria. Each point represents one protein. The x-axis indicates the median ΔNPQ between processing conditions and the y-axis shows Spearman's $r_s$ across paired samples. Dashed lines denote predefined agreement thresholds (|ΔNPQ| ≤ 0.5 and $r_s$ ≥ 0.75). Biomarker performance was classified as uncertain if ≥50% of reference samples were below LLOQ + 0.5 NPQ; otherwise as high, moderate or low according to the predefined bias and correlation criteria.*
